# Longitudinal trends and risk factors for depressed mood among Canadian adults during the first wave of COVID-19

**DOI:** 10.1101/2021.01.21.20245795

**Authors:** Gustavo S. Betini, John P. Hirdes, Rhéda Adekpedjou, Christopher M. Perlman, Nathan Huculak, Paul Hébert

**Affiliations:** School of Public Health and Health Systems, University of Waterloo, Waterloo, ON, Canada; Department de Medicine, Université de Montréal et Centre hospitalier de l’Université de Montréal, Montreal, QC, Canada; Canadian Red Cross

## Abstract

**Background:** The COVID-19 pandemic has raised serious concerns about the mental health impact of people directed and indirectly affected by the virus. Because this is a rapidly evolving situation, our goal was to explore potential risk factors and trends in feelings of anxiety and depression among the general population in Canada over the first five months of the pandemic.

**Methods:** We completed on-line surveys of 3,127 unique individuals representative of the Canadian general population at 4 discreet periods every 6 weeks from April 15^th^ to July 28^th^ 2020. We assessed feelings of anxiety, depression and loss of interest with the interRAI self-reported mood scale using a multivariable generalized estimating equation model to examine factors associated with having a 5+ score on the scale (indicating potentially depressed mood). We also investigated potential longitudinal trends to examine temporal changes in mood scores.

**Results:** More than 30% of participants felt highly anxious, depressed, and disinterested in everyday activities in the first survey (April), but this number decreased to about 20% over 4 months. Feeling lonely, younger age, feeling overwhelmed by one’s health needs, having financial concerns, and living outside of Québec were significantly associated with depressed mood.

**Interpretation:** The prevalence of depressed mood during the pandemic was between 2 and 3 times the pre-pandemic rate (especially among young people), but it can change rapidly in response to social changes. Thus, monitoring of psychological distress among vulnerable groups that may benefit from additional supports should be a priority.

## Introduction

There has been a growing concern that, without focused mitigation efforts, the COVID-19 pandemic has the potential to increase mental health problems worldwide (1–6). In addition to fear of contracting COVID-19, lock-downs, uncertainty, self-isolation and social distancing are disrupting everyday lives, creating personal, social and economic challenges with potential negative psychological effects. This means that the general population can also be adversely affected by mental health consequences of pandemics. International evidence to date clearly suggests that mental health considerations should be taken into account in addition to the physical effects of the virus (7–9,8). For example, quarantine has been reported to cause post-traumatic stress symptoms, confusion, and anger with potentially long lasting effects (7) and depression and anxiety were estimated to be higher among quarantined individuals during the initial stages of COVID-19 in China (8). In addition, risk factors for mental health problems during COVID-19 are reported to include female gender, younger age, presence of chronic and psychiatric illnesses, unemployment, student status, and frequent exposure to social media or news concerning the pandemic (8,14–16,24,25).

Although these studies provide good evidence of the importance of understanding the mental health impact of COVID-19 on the general population, it is still not clear how these negative effects might change with the dynamics of COVID-19 and the changes in the public health policies aiming to contain its spread. This is important because most of these studies were conducted as cross-sectional snapshots at varying time periods of the pandemic, making comparison among studies difficult (8). The objective of this study to examine the mental health impacts of COVID-19 as well as longitudinal changes in the general Canadian population.

## Methods

### Web-based survey

We conducted web-based interviews with the general adult population in Canada from April to July in four discrete surveys, 4-6 weeks apart from each other (Table 1). We used a professional polling company to obtain a sample that was representative of the Canadian population (Table 1) when applying survey weights. Participants were recruited via phone (60%), invitation (25%), social media (5%), offline recruitment (5%), partnerships and campaigns (5%). Among the 3,127 participants, ∼80% were present in the two or more surveys and 1,510 (66%) were present in all surveys (Table 1). Mental health status was assessed with 3 questions from the interRAI self-reported mood scale, which assesses levels of anxiety, depression, and loss of interest (26). The questions were: “In the last 3 days, how often have you felt: **a**. anxious, restless, or uneasy, **b**. sad, depressed, or hopeless and **c**. little interest or pleasure in things you normally enjoy”. Each item has scores ranging from 0 (not present) to 3 (daily), and scores for the three items are summed to create a scale with a value between 0-9. Higher scores representing more frequent and varied mood symptoms (Cronbach’s alpha=0.81). We set a threshold for having substantially depressed mood at 5 or more based on previous analyses that indicate that was a threshold associated with suicide-related ideation in community mental health populations (results available on request). Socio-demographic variables and main concerns before and during the pandemic (e.g., financial concerns, food insecurity levels and loneliness) were also assessed during the interviews (Table 1, 2).

**Table 1.** Profile of the participants in each survey. Number between parenthesis on the sample size column represents new participants added to the survey.

|  | APRIL 15<sup>TH</sup> TO 20<sup>TH</sup> | MAY 6<sup>TH</sup> TO 13<sup>TH</sup> | JUNE 3<sup>RD</sup> TO 9<sup>TH</sup> | JULY 22<sup>TH</sup> TO 28<sup>TH</sup> |
| --- | --- | --- | --- | --- |
| **N** | 2,200 | 2,264 (314) | 2,280 (352) | 2,201 (241) |
| **RECONTACTS IN PREVIOUS SURVEY** | na | 86% | 84% | 87% |
| **GENDER** |  |  |  |  |
| Male | 48% | 48% | 49% | 49% |
| Female | 52% | 52% | 51% | 51% |
| **AGE** |  |  |  |  |
| 18-34 | 27% | 27% | 26% | 24% |
| 35-54 | 34% | 34% | 36% | 37% |
| 55-64 | 17% | 17% | 17% | 17% |
| 65-74 | 12% | 12% | 12% | 12% |
| 75+ | 9% | 9% | 9% | 9% |
| **PROVINCE** |  |  |  |  |
| British Columbia | 14% | 14% | 14% | 14% |
| Alberta | 11% | 11% | 11% | 11% |
| Manitoba/Saskatchewan | 6% | 6% | 7% | 7% |
| Ontario | 38% | 38% | 38% | 38% |
| Quebec | 23% | 23% | 23% | 23% |
| Atlantic | 7% | 7% | 7% | 7% |
| **REGION** |  |  |  |  |
| Quebec | 23% | 23% | 23% | 23% |
| Rest of Canada | 77% | 77% | 77% | 77% |
| **AREA TYPE** |  |  |  |  |
| Urban | 88% | 89% | 90% | 90% |
| Rural | 12% | 11% | 10% | 10% |
| **MOTHER TONGUE** |  |  |  |  |
| French | 21% | 20% | 20% | 20% |
| English | 67% | 66% | 66% | 65% |
| Other languages | 12% | 13% | 14% | 14% |
| **ETHNIC ORIGIN** |  |  |  |  |
| Caucasian (White) | 83% | 81% | 81% | 80% |
| Aboriginal / First Nations | 1% | 1% | 1% | 1% |
| Black | 2% | 2% | 2% | 2% |
| Chinese | 3% | 5% | 5% | 5% |
| Other | 9% | 10% | 10% | 10% |
| **CHILDREN IN THE HOUSEHOLD** |  |  |  |  |
| Yes | 28% | 28% | 28% | 28% |
| No | 72% | 72% | 72% | 72% |
| **LIVING SITUATION** |  |  |  |  |
| Alone | 20% | 20% | 21% | 21% |
| With spouse (partner only) | 32% | 31% | 32% | 32% |
| With spouse / partner and other(s) | 27% | 27% | 26% | 26% |
| With child(ren) (no spouse / partner) | 5% | 5% | 6% | 6% |
| With parent(s) or guardian(s) | 9% | 11% | 10% | 10% |
| With sibling(s) | 1% | 1% | 1% | 1% |
| With other relative(s) | 2% | 2% | 2% | 2% |
| With nonrelative(s) | 3% | 3% | 3% | 3% |
| **VULNERABLE SENIOR** |  |  |  |  |
| Yes | na | 2% | 2% | 2% |
| No | na | 98% | 98% | 98% |
| **EDUCATION** |  |  |  |  |
| Elementary/High School | 33% | 31% | 32% | 30% |
| College | 40% | 41% | 41% | 43% |
| University | 27% | 27% | 27% | 27% |
| **OCCUPATION** |  |  |  |  |
| Office / services / sales | na | 23% | 22% | 23% |
| Manual Worker | 11% | 10% | 10% | 9% |
| Professional | 19% | 19% | 20% | 20% |
| Homemaker | 3% | 4% | 4% | 4% |
| Student | 7% | 7% | 7% | 6% |
| Retired | 27% | 27% | 28% | 28% |
| Unemployed | 5% | 5% | 5% | 4% |
| **HOUSEHOLD INCOME** |  |  |  |  |
| Less than \$40K | 23% | 21% | 22% | 22% |
| \$40K - \$59K | 17% | 18% | 17% | 15% |
| \$60K - \$79K | 15% | 16% | 16% | 15% |
| \$80K - \$99K | 13% | 13% | 14% | 15% |
| \$100K - \$149K | 15% | 15% | 16% | 15% |
| > \$150K | 7% | 7% | 8% | 8% |

**Table 2.** Unadjusted and adjusted odds ratios, 95% confidence interval and P-value for score of 5+ on self-reported mood scale in survey 1 (April 10^th^-15^th^).

|  | Bivariate analysis<br>Unadjusted OR |  |  | Multivariable analysis<br>Adjusted OR |  |  |
| --- | --- | --- | --- | --- | --- | --- |
|  | OR | 95% CI | P-value | OR | 95% CI | P-value |
| **Age (years)** |  |  | **< 0.001** |  |  | **< 0.001** |
| 18-24 vs ≥ 65 | 5.61 | 3.43 to 9.17 | < 0.001 | 2.62 | 1.58 to 4.32 | < 0.001 |
| 25-34 vs ≥ 65 | 3.52 | 2.37 to 5.22 | < 0.001 | 1.96 | 1.21 to 3.17 | 0.006 |
| 35-44 vs ≥ 65 | 3.39 | 2.28 to 5.03 | < 0.001 | 2.67 | 1.73 to 4.14 | < 0.001 |
| 45-64 vs ≥ 65 | 2.31 | 1.69 to 3.15 | < 0.001 | 1.88 | 1.30 to 2.72 | < 0.001 |
| **Gender** |  |  | **0.004** |  |  |  |
| Female vs Male | 1.49 | 1.13 to 1.95 | 0.004 |  |  |  |
| **Province** |  |  | **< 0.001** |  |  | **0.01** |
| Others vs Quebec | 2.1 | 1.49 to 2.94 |  | 1.64 | 1.11 to 2.43 |  |
| **Language** |  |  | **< 0.001** |  |  |  |
| English vs French | 1.89 | 1.33 to 2.69 | < 0.001 |  |  |  |
| Bilingual vs French | 3.21 | 1.59 to 6.47 | 0.001 |  |  |  |
| Others vs French | 1.90 | 1.08 to 3.35 | 0.03 |  |  |  |
| **Education** |  |  | 0.70 |  |  |  |
| College vs Elementary/High school | 0.94 | 0.66 to 1.33 | 0.73 |  |  |  |
| University vs Elementary/High school | 1.07 | 0.77 to 1.47 | 0.69 |  |  |  |
| **Living with** |  |  | **< 0.001** |  |  |  |
| Spouse/partner vs Alone | 0.75 | 0.52 to 1.08 | 0.12 |  |  |  |
| Parent/guardian vs Alone | 1.99 | 1.16 to 3.40 | 0.01 |  |  |  |
| Other relatives vs Alone | 1.23 | 0.86 to 1.76 | 0.25 |  |  |  |
| Non relatives vs Alone | 2.16 | 0.98 to 4.77 | 0.06 |  |  |  |
| **Occupation** |  |  | **< 0.001** |  |  |  |
| Homemade vs Manual worker | 0.94 | 0.41 to 2.12 | 0.88 |  |  |  |
| No answer vs Manual worker | 5.33 | 1.46 to 19.49 | 0.01 |  |  |  |
| Retired vs Manual worker | 0.43 | 0.32 to 0.57 | <0.001 |  |  |  |
| Student vs Manual worker | 1.78 | 1.04 to 3.04 | 0.04 |  |  |  |
| Unemployed vs Manual worker | 1.31 | 0.69 to 2.49 | 0.42 |  |  |  |
| **Income** |  |  | **< 0.001** |  |  |  |
| < \$40,000 vs ≥ \$150,000 | 1.39 | 0.76 to 2.54 | 0.28 | | | |
| \$40,000 - \$59,999 vs ≥ \$150,000 | 1.08 | 0.58 to 2.00 | 0.80 | | | |
| \$60,000 - \$79,999 vs ≥ \$150,000 | 1.57 | 0.83 to 2.97 | 0.16 | | | |
| \$80,000 - \$99,999 vs ≥ \$150,000 | 1.35 | 0.70 to 2.58 | 0.36 | | | |
| \$100,000 - \$149,999 vs ≥ \$150,000 | 0.83 | 0.43 to 1.60 | 0.59 | | | |
| No answer vs ≥ \$150,000 | 0.89 | 0.44 to 1.81 | 0.75 | | | |
| **Ethnicity** |  |  | 0.07 |  |  |  |
| Caucasian vs Other | 1.01 | 0.69 to 1.48 | 0.94 |  |  |  |
| No answer vs Other | 3.49 | 1.17 to 10.43 | 0.03 |  |  |  |
| **Health before Covid19** |  |  | **< 0.001** |  |  |  |
| Fair vs Excellent/good | 1.93 | 1.35 to 2.80 | 0.003 |  |  |  |
|  | OR | 95% CI | P-value | OR | 95% CI | P-value |
| Poor vs Excellent/good | 3.81 | 1.57 to 9.26 | < 0.001 |  |  |  |
| **Health in the past month** |  |  | **< 0.001** |  |  |  |
| Fair vs Excellent/good | 2.73 | 1.98 to 7.36 | < 0.001 |  |  |  |
| Poor vs Excellent/good | 6.15 | 3.01 to 12.52 | < 0.001 |  |  |  |
| **Family overwhelmed by one's health needs** |  |  |  |  |  | < 0.001 |
| Yes vs No | 4.87 | 3.13 to 7.56 | <.001 | 3.92 | 2.39 to 6.41 |  |
| **Feel lonely** |  |  | **< 0.001** |  |  | **< 0.001** |
| Only in specific situations / I do not feel lonely | 2.84 | 1.75 to 4.62 | **< 0.001** | 2.20 | 1.31 to 3.70 | 0.003 |
| Occasionally / I do not feel lonely | 4.74 | 3.1 to 7.22 | **< 0.001** | 3.80 | 2.45 to 5.91 | < 0.001 |
| Daily / I do not feel lonely | 21.31 | 13.66 to 33.26 | **< 0.001** | 16.65 | 10.49 to 26.42 | < 0.001 |
| **Worried about ends meet now** |  |  | **< 0.001** |  |  |  |
| Before and after COVID-19 vs None | 3.37 | 2.45 to 4.64 | < 0.001 | 1.93 | 1.35 to 2.77 | 0.003 |
| Before or after COVID-19 vs None | 2.39 | 1.70 to 3.37 | < 0.001 | 1.69 | 1.11 to 2.58 | 0.02 |
| **Have a way to get food and medication** |  |  | **< 0.001** |  |  |  |
| Yes vs No | 2.77 | 1.65 to 4.66 | **< 0.001** |  |  |  |
| **Feel safe and comfortable in home** |  |  | **< 0.001** |  |  |  |
| Sometimes vs Never | 0.56 | 0.20 to 1.56 | 0.27 |  |  |  |
| Always vs Never | 0.12 | 0.05 to 0.32 | < 0.001 |  |  |  |
| **Have people to count on** |  |  | **< 0.001** |  |  |  |
| Sometimes vs Never | 0.54 | 0.31 to 0.96 | 0.04 |  |  |  |
| Always vs Never | 0.27 | 0.16 to 0.44 | < 0.001 |  |  |  |
| **Can get help if needed** |  |  | **< 0.001** |  |  |  |
| Sometimes vs Never | 0.53 | 0.32 to 0.87 | 0.01 |  |  |  |
| Always vs Never | 0.23 | 0.15 to 0.36 | < 0.001 |  |  |  |

### Statistical analysis

To understand the risk factors associated with the mental health impact of COVID-19, we used a bivariate regression with the percentage of respondents with a 5+ score in the self-reported mood scale as response variable and socio-demographic factors as explanatory variables (Table 1, 2). For simplicity, we only used data from survey 1 for an initial logistic regression model given that the levels of depression, anxiety and loss of interest were stronger at this stage (Figure 1). We then examined a longitudinal interaction between age and survey in a generalized estimating equation model to investigate potential temporal trends in the mental health impact among different age groups. We focused on age because of the great physical health impact that COVID-19 has on older adults (8,18). We did not find a significant interaction between age and survey wave, therefore we presented the model with main effects only. We weighted all analysis using the survey weights to match the sample to population distributions in the latest Statistics Canada census according to gender, age, region, education, mother tongue, living arrangements, and presence of children in the household. We used data from two general population surveys in the Waterloo Region done in 2011 and 2019 (data available upon request) to compare our results with a base level of the same indicators for the general population before the beginning of the COVID-19 pandemic. The baseline level of scores of 5+ on the self-reported mood scale in those surveys ranged between 6.5% in 2011 and 9.7% in 2019, which is comparable to anxiety (6.33%-50.9%, including mild to severe levels) and depression levels (3.6-7.2%) reported in other studies conducted before the pandemic (22). To provide contextual information, daily COVID-19 cases (Figure 1) were obtained from (27) and figures were produced with the *ggplot2* package in R (28).

**Figure 1.**
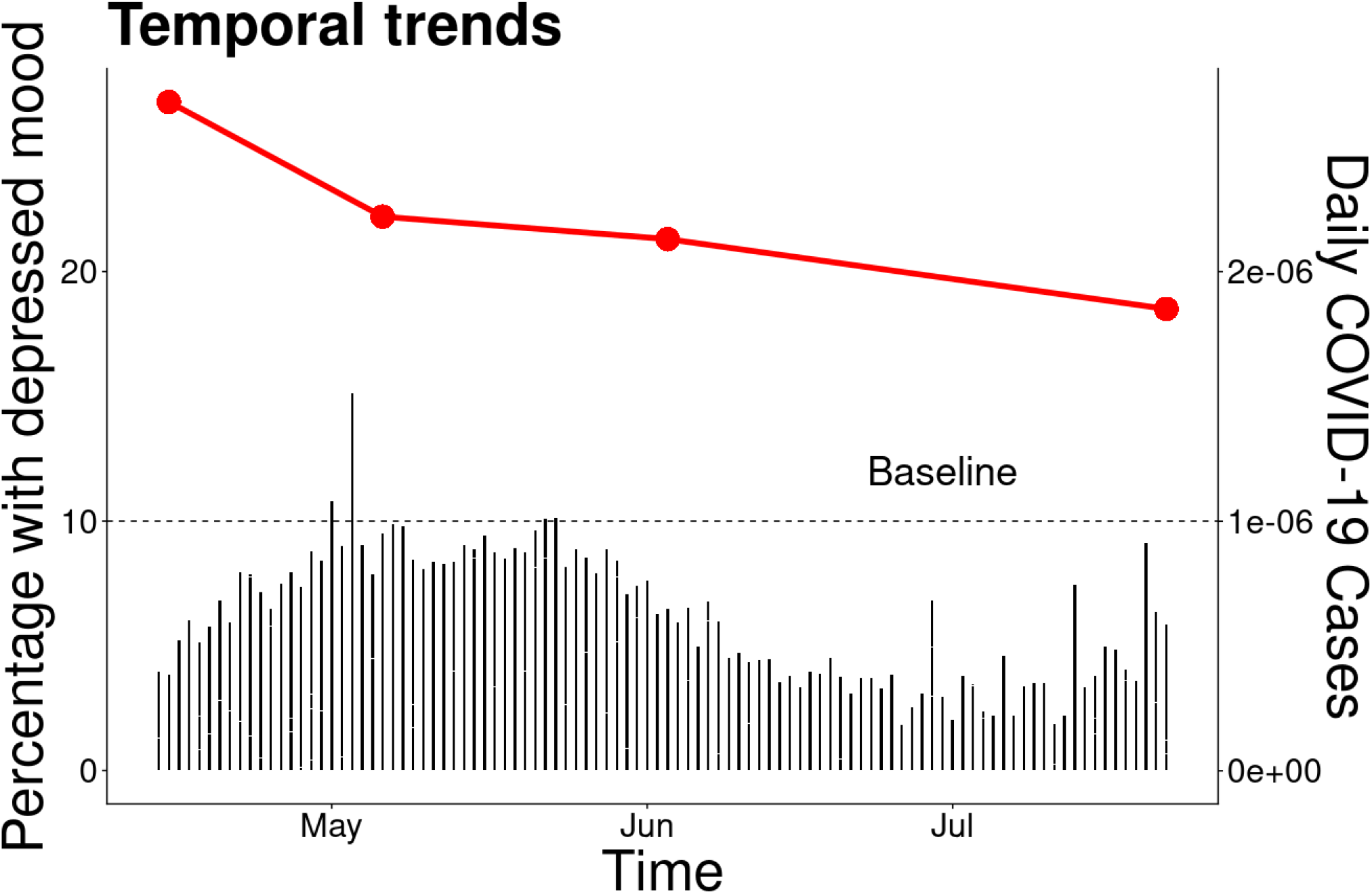
Longitudinal trends in percentage of participants with 5+ score on the self-reported mood scale by study survey.

## Results

We found that up to 44.3% of the participants had substantial level of depressed mood based on indicators of anxiety, depression and loss of interest in the April survey. Only education and ethnicity were not significant risk factors in the bivariate analysis (Table 2). The final multivariable model indicated that age (F_4,2216_ = 7.26, P < 0.001), province (F_1,2219_ = 6.14, P = 0.013), feeling overwhelmed by one’s health needs (F_1,2219_ = 29.56, P = 0.001), loneliness (F_1,2217_ = 52.37, P < 0.0001), and financial concerns (F_1,2218_ = 7.13, P < 0.001) were significantly associated with depressed mood (F_11_,_2209_ = 56.74, P < 0 .001; c-statistics = 0.836). The odds of having a depressed mood were 2.62 times higher in young (18-24) than older adults (65+; 95% confidence interval [CI] 1.58-4.32, P < 0.001), 1.64 times higher in people from other provinces compared to Québec (95% CI 1.11-2.43, P = 0.013), 3.92 times higher in people who felt overwhelmed by their health needs (95% CI 2.39-6.41, P < 0.001), 16.65 times higher in people who felt lonely daily compared to those that did not feel lonely (95% CI 10.49-26.42, P < 0.0001) and 1.93 times higher in people that had financial concerns before and after the pandemic than people without financial concerns (95% CI 1.35-2.77, P < 0.001; Table 2, Figure 1). We found a significant temporal trend in the generalized estimating equation model, suggesting a decrease in the odds of depressed mood over time compared with the initial stage of the survey done in April 2020 (Table 3, Figure 1).

**Table 3.** Parameter estimates, odds ratio (OR), 95% confidence interval (95% CI) and P-value for the longitudinal multivariable generalized estimating equation model for self-report mood scale score of 5+ over four study waves.

| Parameter | Estimate | 95% CI |  | Z | P-value | Adjusted OR | 95% CI |  |
| --- | --- | --- | --- | --- | --- | --- | --- | --- |
| **Intercept** | -2.761 | -3.126 | -2.398 | -14.87 | < 0.001 |  |  |  |
| **Age group (ref=18-24)** |  |  |  |  |  |  |  |  |
| 25-34 | 0.12 | -0.20 | 0.45 | 0.74 | 0.45 | 1.13 | 0.82 | 1.56 |
| 35-44 | -0.23 | -0.58 | 0.12 | -1.28 | 0.20 | 0.79 | 0.56 | 1.13 |
| 45-64 | -0.25 | -0.54 | 0.03 | -1.73 | 0.08 | 0.78 | 0.58 | 1.03 |
| 65+ | -0.79 | -1.07 | -0.51 | -5.56 | < 0.001 | 0.45 | 0.34 | 0.60 |
| **Province (ref=Quebec)** |  |  |  |  |  |  |  |  |
| Others | 0.26 | 0.05 | 0.46 | 2.46 | 0.01 | 1.30 | 1.05 | 1.59 |
| **Health Status (ref=no concerns)** |  |  |  |  |  |  |  |  |
| Overwhelmed by one's health needs | 1.33 | 1.05 | 1.62 | 9.16 | < 0.001 | 3.80 | 2.85 | 5.05 |
| **Loneliness (ref=not lonely)** |  |  |  |  |  |  |  |  |
| Lonely only in specific situations | 0.74 | 0.45 | 1.03 | 4.94 | < 0.001 | 2.09 | 1.56 | 2.80 |
| Lonely occasionally | 1.39 | 1.14 | 1.64 | 10.83 | < 0.001 | 4.02 | 3.13 | 5.17 |
| Lonely daily | 2.96 | 2.69 | 3.22 | 22.11 | < 0.001 | 19.29 | 14.84 | 25.07 |
| **Financial status (ref=no worries)** |  |  |  |  |  |  |  |  |
| Financial worry before and after COVID-19 | 0.69 | 0.48 | 0.89 | 6.56 | < 0.001 | 2.00 | 1.62 | 2.45 |
| Financial worry before or after COVID-19 | 0.62 | 0.38 | 0.86 | 5.09 | < 0.001 | 1.86 | 1.46 | 2.35 |
| **Study Wave (ref=Survey 1)** |  |  |  |  |  |  |  |  |
| Survey 2 | -0.23 | -0.46 | 0.01 | -1.89 | 0.06 | 0.80 | 0.63 | 1.01 |
| Survey 3 | -0.27 | -0.52 | -0.03 | -2.20 | 0.03 | 0.76 | 0.60 | 0.97 |
| Survey 4 | -0.32 | -0.57 | -0.07 | -2.52 | 0.01 | 0.72 | 0.56 | 0.93 |

## Interpretation

Our study provided a longitudinal view of the mental health impact of COVID-19 on about 3,000 participants followed over a 4-month period. The impact was most pronounced on the mental health of younger Canadians and those who reported feeling lonely. In addition, the odds of serious mood disturbance were strongest at the beginning of the pandemic (April), with a rapidly decrease from April to July. However, the absolute levels in July were still 2 times higher compared to the pre-pandemic levels. Although there appears to be potential for resilience and fast recovery in part of the population, the absence of complete recovery could result in even higher levels of anxiety and depression during new waves of the pandemic.

Our results are in line with other studies on the mental health effects of COVID-19, both in the strength of the association and key risk factors (8,11,13,14,23). A meta-analysis found that during the pandemic, prevalence of depression symptoms was 33.7% (95% confidence interval 27.5-40.6) and 31.9% for anxiety (95% CI 27.5-36.7; 16). As in our study, these levels were, in average, higher than the pre-pandemic levels we noted in earlier studies based on our measure (between 6.5 and 9.7%) as well as comparable rates reported by others; (8). Other longitudinal studies have reported a mix of results, with small increases in feelings of depression and decreased anxiety (29) or no trend overall (30). Our results showed roughly a 30% decrease in the odds of disturbed mood spanned a 4-month period, whereas other longitudinal studies investigated trends over a much shorter time period (29,30). Some of the discrepancies reported in the published literature could be explained by the phase of the pandemic when the study was conducted.

We also observed that young respondents were among the most affected groups, even after controlling for potential confounding factors such as employment status, gender, health, and economic status. This may reflect a true age-group difference in the COVID-19 experience, but it may also reflect generational differences in comfort related to reporting mental health symptoms. Student status and gender have been identified as risk factors in other studies (8), but these were only significant in our bivariate model. Although student status was strongly associated with age, which may explain it not persisting as a predictor of mood disturbance in the multivariable model, it is less clear why gender was not a significant risk factor when we controlled for other sociodemographic variables.

Although the levels of anxiety and depression reported here are in line with the published literature (8,18), COVID-19 happened during a time of changing public sentiment about global political, economic, and climate stability with increased focus on natural disasters like wild fires (e.g., bushfires in Australia and California), and an increase in public protest related to racial equality. Thus, it is unclear from our results whether the pandemic was the primary driver of changes in mood that we observed or it was only one of the many contributors, potentially acting as an amplifier of these other source of stress (31).

We observed clear trends on mental health indicators on a period of 4 months. However, we still do not know what the long-term consequences of COVID-19 will be nor what policies will successfully mitigate its mental health impacts. Information on the long-term impact of past pandemics, such as the Spanish Flu, is scarce. However, some studies reported that people developed psychiatric disorders several years after the 2003 SARS-CoV-1 pandemic (32). Moreover, studies on natural disasters, such as hurricane, fires and earthquake also point to long-term effects where lifetime post-traumatic stress disorder rates can be up to 40% higher in disaster survivors compared to controls (33,34). Given the substantially elevated levels of distressed mood compared with pre-COVID-19 levels, it is important to monitor whether long-term mental health effects persist in the general population.

## Limitations

Our overall response rate was in the 35% range and the assembled sample was representative of Canadians. In addition, by repeatedly sampling more than 80% of the same individuals over time, we are confident that temporal trends were accurately measured. Despite this, vulnerable groups may not have been well represented. For instance, older socially isolated adults, persons in facility-based settings (e.g., long-term care), and other marginalized groups may not have internet access or may not be able to participate because of other barriers (35–37). In addition, depressive symptoms tend to decrease with age (38,39). Thus, the absolute values of our mental health indicators might be biased, specifically among older adults (65+), despite the fact that socio-demographic factors were weighted in the statistical analysis. Fortunately, non-response to surveys does not substantially harm the ability to estimate associations among variables including to investigate temporal trends (35). Another limitation in our web-survey approach is the limited subsample sizes of minority groups who may be deferentially affected by mental health concerns. Future efforts to examine the impact of COVID-19 on race and ethnicity should over-sample minority groups to allow for adequate subsample sizes.

## Conclusions

In our survey, we showed that the pandemic increased feelings of anxiety, depression, and loss of interest symptoms 2 to 3-fold, especially in young people. We also documented that these changes can rapidly decrease in a short period of time, likely related to external social trends such as implementation of broad social policies related to epidemic control, communications from media, health experts, and political leadership. Future studies should focus not only on the description of the mental health consequences, but also in establishing evidence of possible causal relationships between the dynamic of the disease, public health policies and mental health indicators. For example, it would be important to tease apart the effects of fear of disease, subjective and objective aspects of social isolation, economic uncertainty, other challenges to mental health.

## Data Availability

Data is available upon request.

## Acknowledgements

We thank Wendy Hepburn, Mary Bartram, and Raquel Betini for comments on early versions of this manuscript. This project was funded by the Canadian Red Cross and survey conducted by Leger. The Canadian Red Cross provided all data to the investigative team without constraints.

Disclaimer: Dr. Paul Hébert and Nathan Huculak are employed by the Canadian Red Cross. Opinions expressed herein are solely those of the investigators. They do not necessarily reflect views or positions held by the Canadian Red Cross.

## Authors Contribution

JPH, CMP, NH, and PH designed the study; JPH and RA analyzed the data; GSB wrote the first draft of the manuscript. All authors discussed the ideas and commented on subsequent drafts of the manuscript.

## Notes

**Funding:** supported by a contribution from the Canadian Red Cross

### Competing Interest Statement

The authors have declared no competing interest.

### Funding Statement

This study was supported by a contribution from the Canadian Red Cross

### Author Declarations

We obtained ethics clearance from the University of Waterloo's Research Ethics Committee (ORE#42932)

